# A Comparative Genomic Analysis of Left- and Right-Sided Colon Cancer Using Real-World Data from the AACR Project GENIE BPC Dataset

**DOI:** 10.1101/2025.06.30.25330590

**Authors:** Younggwang Kim, Min Ki Kim, Sanghun Lee

## Abstract

Right-sided colon cancer (RCC) and left-sided colon cancer (LCC) exhibit distinct clinical and molecular characteristics, influencing prognosis and therapeutic strategies. This study analyzed 750 patients with histologically confirmed adenocarcinoma (RCC: 387, LCC: 363) from the AACR Project Genomics Evidence Neoplasia Information Exchange (GENIE) Biopharma Collaborative (BPC) dataset. Tumor mutation burden (TMB) was significantly higher in RCC (6.65 ± 11.3) than in LCC (3.17 ± 4.35, adjusted *p* = 3.12 × 10^-32^). After filtering for functionally significant mutations, RCC exhibited enrichment for *BRAF* (23.1% vs. 6.7%, adjusted *p* = 1.64 × 10^-8^), *KMT2D* (8.6% vs. 3.2%, adjusted *p* = 9.27 × 10^-3^), and *SMAD4* (13.1% vs. 7.3%, adjusted *p* = 2.97 × 10^-2^) mutations, while LCC demonstrated a higher frequency of *TP53* (40.6% vs. 31.8%, adjusted *p* = 2.97 × 10^-2^). Multivariate Cox regression analysis showed significantly poorer overall survival (OS) in RCC than in LCC (HR: 1.30, 95% CI: 1.02–1.66, *p* = 0.033). *KRAS* mutations were identified as an independent predictor of worse OS in RCC (HR: 1.68, 95% CI: 1.06–2.70, *p* = 0.027), while *BRAF* mutations were associated with poorer OS in LCC (HR: 1.58, 95% CI: 1.05–2.37, *p* = 0.028).

## Introduction

Colon cancer (CC) is a leading cause of cancer-related deaths in developed countries (1). It arises from the epithelial lining of the colon and can occur on either the right (proximal) or left (distal) sections of the colon. Tumor location plays a critical role in disease progression and overall survival outcomes (2). Right-sided colon cancer (RCC) and left-sided colon cancer (LCC) are distinct entities with differing epidemiological, clinicopathological, and molecular characteristics. These distinctions are driven by variations in gene expression profiles, with more than 1,000 genes exhibiting differential expression between RCC and LCC (2). Specifically, 165 genes exhibit over a two-fold difference, and 49 genes show over a three-fold difference, reflecting intrinsic biological differences established during embryonic development and maintained throughout postnatal life (2).

The genomic landscapes of RCC and LCC further highlight their divergence. RCC is frequently associated with microsatellite instability (MSI)-high tumors and CpG island methylator phenotype (CIMP) positivity, whereas LCC is predominantly characterized by chromosomal instability (CIN-high) (2, 3). These molecular differences are reflected in histological features: RCC commonly presents with flat morphology, poor differentiation, and mucinous features, which often lead to delayed detection during colonoscopy and advanced tumor stages at diagnosis (4). Conversely, LCC typically exhibits polypoid morphology, making it more amenable to early detection (5). These differences extend to metastatic patterns, with RCC more frequently associated with peritoneal carcinomatosis, while LCC tends to metastasize to the liver and lungs (6). These metastatic tendencies are influenced by the anatomical, vascular, and molecular variations driving tumor progression in each type.

Understanding the clinical, molecular, and histological differences between RCC and LCC is critical for tailoring therapeutic strategies and improving prognostic accuracy. While substantial progress has been made in characterizing these distinctions, many prior studies were limited by small sample sizes and a lack of integrative analysis of both clinical and genomic data (7–10). These issues often resulted in fragmented or conflicting conclusions regarding the molecular and clinical variability between RCC and LCC. To address these challenges, we utilized real-world data from the American Association for Cancer Research (AACR) Project Genomics Evidence Neoplasia Information Exchange (GENIE) Biopharma Collaborative (BPC) dataset, a platform integrating large-scale genomic and clinical datasets from diverse cohorts (11). This resource overcomes many of the limitations of traditional studies, providing a robust foundation for high-resolution analysis.

In this study, we investigate the key clinical, molecular, and mutational differences between RCC and LCC using the GENIE BPC dataset. Our analysis aimed to elucidate how these variations influence clinical presentations, including metastatic patterns and patient outcomes. By utilizing this comprehensive dataset, we hope to provide novel insights into the biological variability between LCC and RCC.

## Materials and methods

### Study Population and Inclusion Criteria

Data of patients with CC were collected from the GENIE BPC CRC v2.0-public dataset. These data were provided by multiple institutions, including Dana-Farber Cancer Institute (DFCI), Memorial Sloan Kettering Cancer Center (MSKCC), Princess Margaret Cancer Centre - University Health Network (UHN), and Vanderbilt-Ingram Cancer Center (VICC). Tumors were classified as LCC (descending colon [C18.5], sigmoid colon [C18.6], or rectosigmoid junction [C18.7]) or RCC (cecum [C18.0], ascending colon [C18.2], hepatic flexure [C18.3], or transverse colon [C18.4]) based on site-specific ICD-O-3 codes. Patients aged 18 or older at diagnosis with at least two years of follow-up data were included in the study, and those with ambiguous tumor locations or histologies other than adenocarcinoma (e.g., mucinous or signet ring cell adenocarcinoma) were excluded (12).

Demographic data, including sex, age at diagnosis, race, and ethnicity, and disease data, including tumor stage at initial diagnosis, tumor grade, presence of distant metastases, lymph node involvement, carcinoembryonic antigen (CEA) levels at diagnosis, MSI status, and treatment regimens, were collected from pathology, radiology, and oncology reports using a structured framework. All data were collected under each ethical approval from each institution, and all patients provided informed consent in accordance with the Declaration of Helsinki.

### Genomic analysis

Next-generation sequencing (NGS) data had been generated to identify single-nucleotide variants and small insertions/deletions. We used genetic data before and after filtering for functionally significant mutations using a PolyPhen score >0.85 and a SIFT score <0.05. Our analysis also focused on frequent genetic alterations in CC, including *TP53*, *KRAS*, *PIK3CA*, *SMAD4*, *BRAF*, *FBXW7*, *ATM*, and *KMT2D*. Tumor mutation burden (TMB) had been provided as the number of genes with non-synonymous mutations per sample. We analyzed these genetic alterations according to tumor laterality and clinical outcomes.

### Statistical analysis

Descriptive statistics were used to summarize baseline clinical and genetic characteristics. Continuous variables, such as age at diagnosis and overall survival (OS), were presented as mean and standard deviation (SD), whereas categorical variables, such as tumor laterality, genetic alterations, and treatment regimens, were reported as counts and percentages. Differences in clinical and genetic characteristics between LCC and RCC were assessed using chi-squared tests or Fisher’s exact tests, as appropriate. Survival analyses were conducted to evaluate OS, defined as the time from diagnosis to death or last follow-up. Kaplan–Meier survival curves were generated to compare OS between groups, and log-rank tests were used to assess statistical significance. Subgroup analyses were conducted to evaluate differences in OS based on tumor laterality, sex, age group (≥ 65 years vs. <65 years), tumor stage at diagnosis (stage IV vs. stages I–III), CEA levels at diagnosis (>5 ng/mL vs. ≤ 5 ng/mL), TMB, and the presence of somatic mutations. To identify independent predictors of OS, multivariate Cox proportional hazards regression models were constructed. The models incorporated tumor laterality, sex, age at diagnosis, tumor stage at diagnosis, CEA levels at diagnosis, TMB, and the presence of somatic mutations in key genes associated with CC. Hazard ratios (HRs) with 95% confidence intervals (CIs) were calculated for each variable to quantify the strength of associations. The proportional hazards assumption was verified using Schoenfeld residuals, and no significant violations were observed. All statistical analyses were performed using R version 4.3.1. Kaplan–Meier survival analyses and Cox proportional hazards regression models were conducted using the “survival” and “survminer” packages; Oncoplot analyses and visualizations were carried out using the “maftools” and “ggplot2” packages.

## Results

### Data Filtering, Annotation, and Grouping Strategy

Fig. 1 illustrates the workflow, outlining the process of patient selection from the AACR Project GENIE BPC database. Initially, 1,485 patients with CC were selected; the cohort was subsequently narrowed to 1,217 patients diagnosed with histologically confirmed adenocarcinoma. After excluding patients younger than 18 at diagnosis and those with insufficient follow-up periods (<2 years), ambiguous tumor locations, or histologies other than adenocarcinoma, the final cohort included 750 patients with CC. These were stratified into LCC (n = 363) and RCC (n = 387) groups.

**Figure 1.**
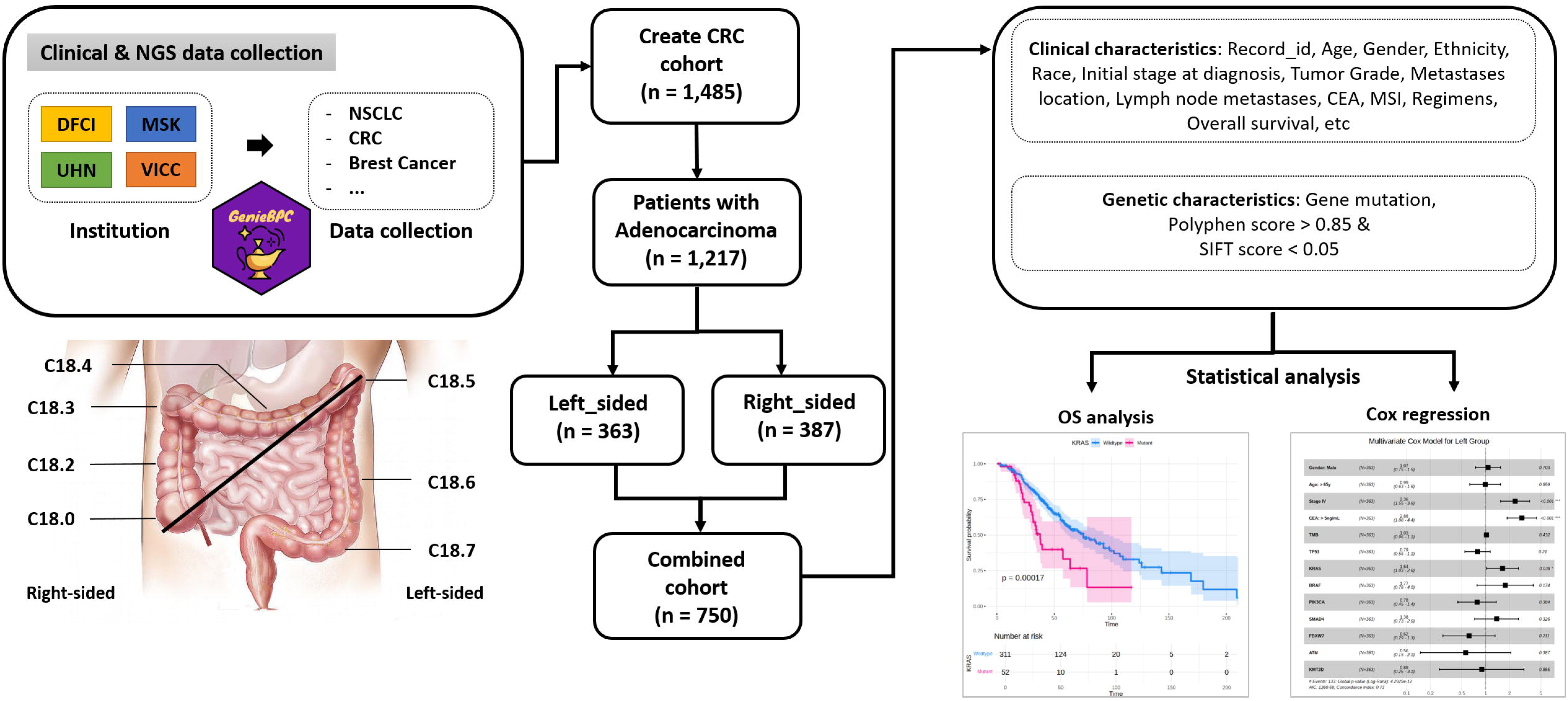
Workflow of patient selection, data annotation, and analysis framework. The flowchart outlines the construction of the colorectal cancer (CRC) cohort from the AACR Project GENIE BPC dataset. After applying clinical filters, 750 patients with histologically confirmed adenocarcinoma were selected and classified into left-sided (n = 363) and right-sided (n = 387) groups based on ICD-O-3 codes. Clinical variables and annotated genomic features (PolyPhen > 0.85, SIFT < 0.05) were integrated for survival and regression analysis.

The data workflow captured a comprehensive array of clinical and genomic parameters essential for analyzing differences between LCC and RCC. Key clinical metrics, such as tumor stage, CEA levels, and MSI status, were meticulously curated from the dataset. Additionally, genomic profiling leveraged advanced annotation tools such as PolyPhen and SIFT, allowing for the identification of mutations with high pathogenic potential. This systematic integration of clinical and molecular data provides a robust foundation for distinguishing the biological and clinical characteristics unique to LCC and RCC, paving the way for insights into their divergent progression patterns.

### Clinical and demographic characteristics

The clinical and demographic characteristics of the cohort revealed several significant differences between the groups (Table. 1). Patients with RCC were significantly older, on average, compared to those with LCC (58.7 ± 13.1 years vs. 52.9 ± 11.7 years, *p* = 3.39×10⁻¹⁰), and the proportion of individuals aged 65 years or older was markedly higher in the RCC group (36.2% vs. 17.6%). Disease stage at diagnosis differed significantly, with stage IV being more prevalent in LCC (55.6%) compared to RCC (43.9%, *p* = 1.12×10⁻⁴). Tumor location distributions demonstrated further contrasts, with the majority of LCC cases located in the sigmoid colon (C18.7, 76.9%), whereas RCC cases were more evenly distributed, predominantly involving the cecum (C18.0, 44.2%) and ascending colon (C18.2, 29.7%). Metastatic patterns, such as liver metastases, showed higher prevalence in LCC (86.6%) compared to RCC (68.2%); however, the difference approached but did not reach statistical significance (*p* = 0.0514). Therapeutic interventions, including chemotherapy and targeted agents, were similar across groups, although RCC had a slightly higher use of chemotherapy (73.1% vs. 63.6%).

**Table 1.**
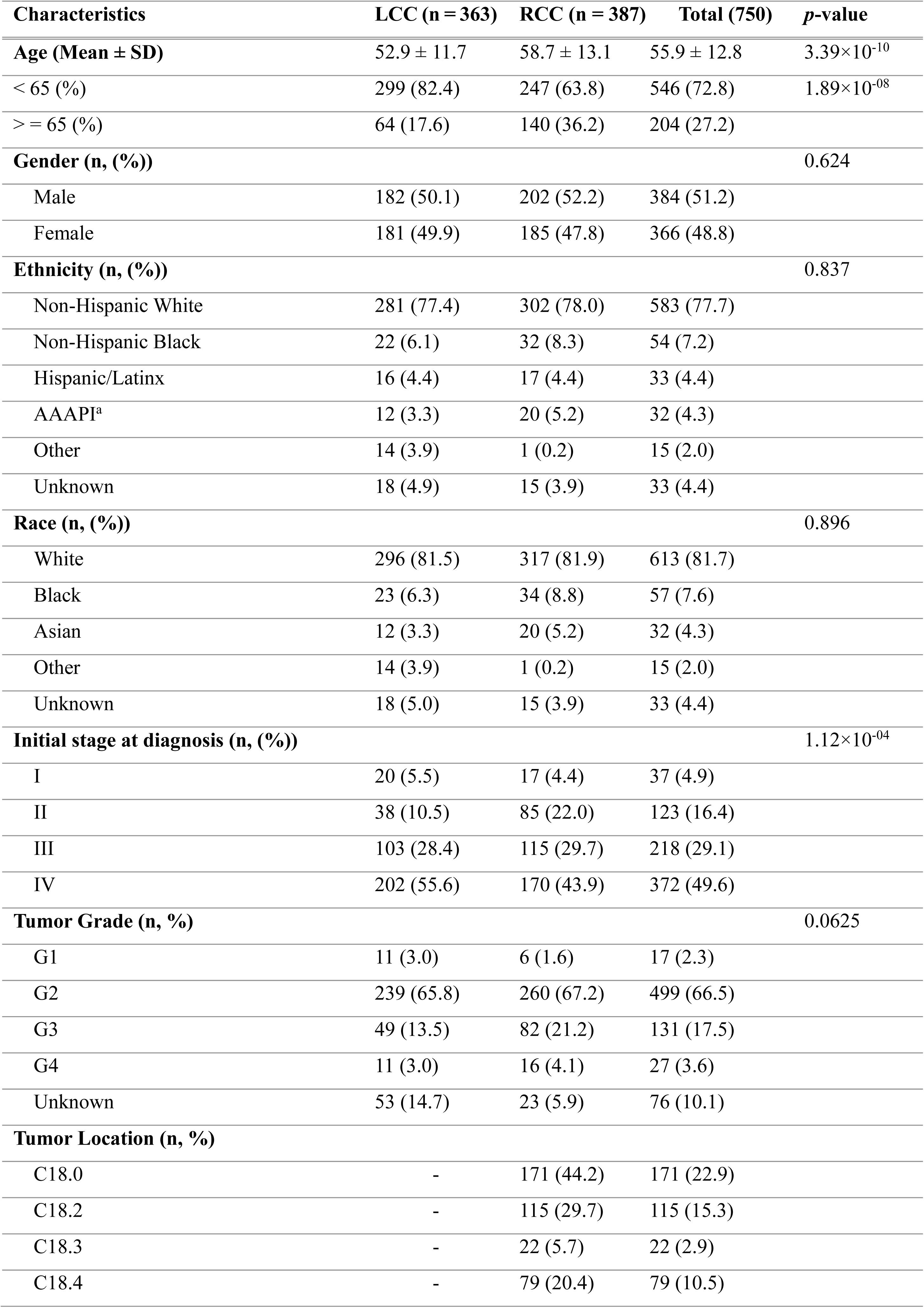
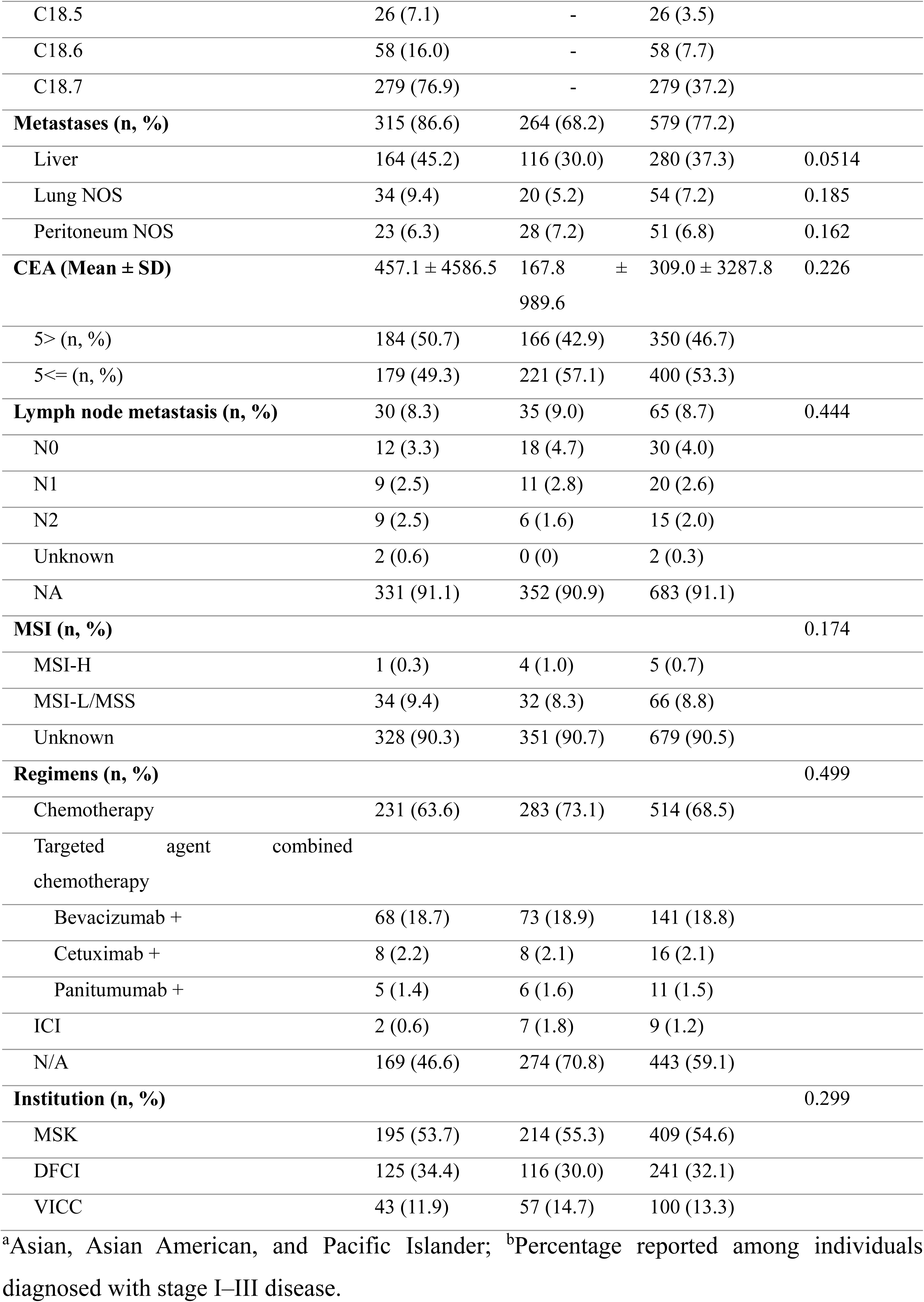
Demographic and Clinical Characteristics of Patients with LCC and RCC.

| Characteristics | LCC (n = 363) | RCC (n = 387) | Total (750) | *p*-value |
| --- | --- | --- | --- | --- |
| **Age (Mean ± SD)** | 52.9 ± 11.7 | 58.7 ± 13.1 | 55.9 ± 12.8 | 3.39×10<sup>-10</sup> |
| < 65 (%) | 299 (82.4) | 247 (63.8) | 546 (72.8) | 1.89×10<sup>-08</sup> |
| > = 65 (%) | 64 (17.6) | 140 (36.2) | 204 (27.2) |  |
| **Gender (n, (%))** |  |  |  | 0.624 |
| Male | 182 (50.1) | 202 (52.2) | 384 (51.2) |  |
| Female | 181 (49.9) | 185 (47.8) | 366 (48.8) |  |
| **Ethnicity (n, (%))** |  |  |  | 0.837 |
| Non-Hispanic White | 281 (77.4) | 302 (78.0) | 583 (77.7) |  |
| Non-Hispanic Black | 22 (6.1) | 32 (8.3) | 54 (7.2) |  |
| Hispanic/Latinx | 16 (4.4) | 17 (4.4) | 33 (4.4) |  |
| AAAPI<sup>a</sup> | 12 (3.3) | 20 (5.2) | 32 (4.3) |  |
| Other | 14 (3.9) | 1 (0.2) | 15 (2.0) |  |
| Unknown | 18 (4.9) | 15 (3.9) | 33 (4.4) |  |
| **Race (n, (%))** |  |  |  | 0.896 |
| White | 296 (81.5) | 317 (81.9) | 613 (81.7) |  |
| Black | 23 (6.3) | 34 (8.8) | 57 (7.6) |  |
| Asian | 12 (3.3) | 20 (5.2) | 32 (4.3) |  |
| Other | 14 (3.9) | 1 (0.2) | 15 (2.0) |  |
| Unknown | 18 (5.0) | 15 (3.9) | 33 (4.4) |  |
| **Initial stage at diagnosis (n, (%))** |  |  |  | 1.12×10<sup>-04</sup> |
| I | 20 (5.5) | 17 (4.4) | 37 (4.9) |  |
| II | 38 (10.5) | 85 (22.0) | 123 (16.4) |  |
| III | 103 (28.4) | 115 (29.7) | 218 (29.1) |  |
| IV | 202 (55.6) | 170 (43.9) | 372 (49.6) |  |
| **Tumor Grade (n, %)** |  |  |  | 0.0625 |
| G1 | 11 (3.0) | 6 (1.6) | 17 (2.3) |  |
| G2 | 239 (65.8) | 260 (67.2) | 499 (66.5) |  |
| G3 | 49 (13.5) | 82 (21.2) | 131 (17.5) |  |
| G4 | 11 (3.0) | 16 (4.1) | 27 (3.6) |  |
| Unknown | 53 (14.7) | 23 (5.9) | 76 (10.1) |  |
| **Tumor Location (n, %)** |  |  |  |  |
| C18.0 | - | 171 (44.2) | 171 (22.9) |  |
| C18.2 | - | 115 (29.7) | 115 (15.3) |  |
| C18.3 | - | 22 (5.7) | 22 (2.9) |  |
| C18.4 | - | 79 (20.4) | 79 (10.5) |  |
| C18.5 | 26 (7.1) | - | 26 (3.5) |  |
| C18.6 | 58 (16.0) | - | 58 (7.7) |  |
| C18.7 | 279 (76.9) | - | 279 (37.2) |  |
| **Metastases (n, %)** | 315 (86.6) | 264 (68.2) | 579 (77.2) |  |
| Liver | 164 (45.2) | 116 (30.0) | 280 (37.3) | 0.0514 |
| Lung NOS | 34 (9.4) | 20 (5.2) | 54 (7.2) | 0.185 |
| Peritoneum NOS | 23 (6.3) | 28 (7.2) | 51 (6.8) | 0.162 |
| **CEA (Mean ± SD)** | 457.1 ± 4586.5 | 167.8 ± 989.6 | 309.0 ± 3287.8 | 0.226 |
| 5> (n, %) | 184 (50.7) | 166 (42.9) | 350 (46.7) |  |
| 5≤ (n, %) | 179 (49.3) | 221 (57.1) | 400 (53.3) |  |
| **Lymph node metastasis (n, %)** | 30 (8.3) | 35 (9.0) | 65 (8.7) | 0.444 |
| N0 | 12 (3.3) | 18 (4.7) | 30 (4.0) |  |
| N1 | 9 (2.5) | 11 (2.8) | 20 (2.6) |  |
| N2 | 9 (2.5) | 6 (1.6) | 15 (2.0) |  |
| Unknown | 2 (0.6) | 0 (0) | 2 (0.3) |  |
| NA | 331 (91.1) | 352 (90.9) | 683 (91.1) |  |
| **MSI (n, %)** |  |  |  | 0.174 |
| MSI-H | 1 (0.3) | 4 (1.0) | 5 (0.7) |  |
| MSI-L/MSS | 34 (9.4) | 32 (8.3) | 66 (8.8) |  |
| Unknown | 328 (90.3) | 351 (90.7) | 679 (90.5) |  |
| **Regimens (n, %)** |  |  |  | 0.499 |
| Chemotherapy | 231 (63.6) | 283 (73.1) | 514 (68.5) |  |
| Targeted agent combined chemotherapy |  |  |  |  |
| Bevacizumab + | 68 (18.7) | 73 (18.9) | 141 (18.8) |  |
| Cetuximab + | 8 (2.2) | 8 (2.1) | 16 (2.1) |  |
| Panitumumab + | 5 (1.4) | 6 (1.6) | 11 (1.5) |  |
| ICI | 2 (0.6) | 7 (1.8) | 9 (1.2) |  |
| N/A | 169 (46.6) | 274 (70.8) | 443 (59.1) |  |
| **Institution (n, %)** |  |  |  | 0.299 |
| MSK | 195 (53.7) | 214 (55.3) | 409 (54.6) |  |
| DFCI | 125 (34.4) | 116 (30.0) | 241 (32.1) |  |
| VICC | 43 (11.9) | 57 (14.7) | 100 (13.3) |  |

<sup>a</sup>Asian, Asian American, and Pacific Islander; <sup>b</sup>Percentage reported among individuals diagnosed with stage I–III disease.

### Mutation profiles

The oncoplot analysis provides a comprehensive view of the mutational landscape before and after applying filtering criteria based on PolyPhen and SIFT (Fig. 2). Prior to filtering, the most frequently mutated genes were *APC* (72%), *TP53* (67%), *KRAS* (45%), *PIK3CA* (27%), *BRAF* (15%), *KMT2D* (15%), *SMAD4* (15%), and *FBXW7* (15%). However, after applying functional annotation filtering, *TP53* (36%) became the most frequently ranked mutation, while the frequencies of other mutations were adjusted: *PIK3CA* (17%), *KRAS* (16%), *BRAF* (15%), *SMAD4* (10%), *FBXW7* (8%), and *KMT2D* (6%). Notably, *APC* mutations, initially the most dominant, were deprioritized due to lower predicted pathogenicity scores, highlighting the importance of functional filtering in refining biologically significant alterations.

**Figure 2.**
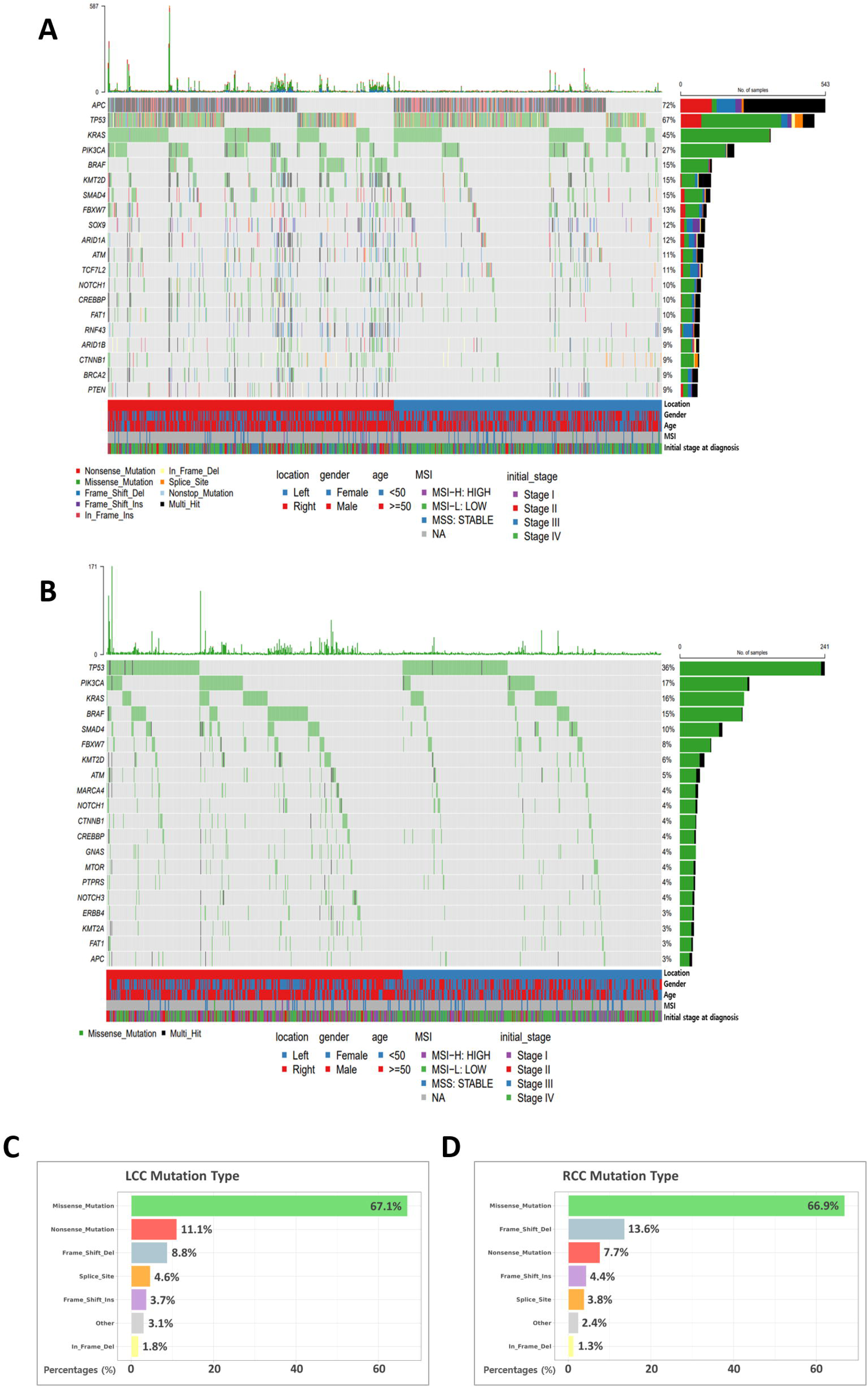
Mutational landscape and mutation type distribution in left-sided and right-sided colorectal cancer. (A) Oncoplot depicting the most frequently mutated genes in the full colorectal cancer cohort prior to functional filtering. *APC*, *TP53*, *KRAS*, and *PIK3CA* were among the most common alterations. (B) Functionally annotated oncoplot highlighting mutations with high predicted pathogenicity based on PolyPhen and SIFT scores. *TP53*, *PIK3CA*, *KRAS*, and *BRAF* were most frequently retained after filtering. (C) Distribution of mutation types in the left-sided colorectal cancer (LCC) group, with missense mutations accounting for the majority. (D) Mutation type distribution in the right-sided colorectal cancer (RCC) group, also dominated by missense mutations. Differences in the proportions of frameshift deletions and nonsense mutations suggest distinct mutational mechanisms between LCC and RCC.

Differences in mutation types between LCC and RCC are evident in the mutation type distributions (Fig. 2C and D). Missense mutations were the most common in both groups, accounting for 67.1% in LCC and 66.9% in RCC. Frameshift deletions, however, were more frequent in RCC (13.6%) compared to LCC (8.8%), while nonsense mutations occurred more frequently in LCC (11.1%) than in RCC (7.7%). These distinct patterns in mutation types suggest differing mutational processes between LCC and RCC

### Comparison of mutation burden and key genetic alterations

To comprehensively compare the molecular differences between LCC and RCC, TMB and key genetic alterations were analyzed before and after applying PolyPhen and SIFT filtering criteria (Table. 2). In the unfiltered analysis, TMB was significantly higher in RCC (6.87 ± 12.1) compared to LCC (3.45 ± 4.39, adjusted *p* = 1.61 × 10⁻³⁰). Among genetic mutations, *APC* mutations were more prevalent in LCC (81.5%) than in RCC (67.2%, adjusted *p* = 1.57 × 10⁻⁵). *TP53* mutations were also more frequent in LCC (73.6%) than in RCC (62.5%, adjusted *p* = 1.54 × 10⁻³), whereas *KRAS* (50.1% vs. 40.2%, adjusted *p* = 7.31 × 10⁻³) and *PIK3CA* (32.6% vs. 20.9%, adjusted *p* = 6.3 × 10⁻⁵) mutations were enriched in RCC. *BRAF* (23.8% vs. 6.6%, adjusted *p* = 3.73 × 10⁻¹⁰) and *KMT2D* (22.5% vs. 8.3%, adjusted *p* = 2.47 × 10⁻⁷) mutations were significantly more frequent in RCC than in LCC, while *SMAD4* mutations were also higher in RCC (19.9%) compared to LCC (10.5%, adjusted *p* = 5.13 × 10⁻⁴). After applying PolyPhen and SIFT filtering criteria, the distribution of mutations changed significantly. TMB remained higher in RCC (6.65 ± 11.3) than in LCC (3.17 ± 4.35, adjusted p = 3.12 × 10⁻³²). *TP53* became the most frequently ranked mutation overall (40.6% in LCC, 31.8% in RCC, adjusted *p* = 2.97 × 10⁻²), while *KRAS* mutations were more balanced between the two groups (16.5% in LCC, 15.0% in RCC, *p* = 0.602). *BRAF* remained significantly enriched in RCC (23.1% vs. 6.7%, adjusted *p* = 1.64 × 10⁻⁸), as did *KMT2D* (8.6% vs. 3.2%, adjusted *p* = 9.27 × 10⁻³) and *SMAD4* (13.1% vs. 7.3%, adjusted *p* = 2.97 × 10⁻²). *PIK3CA* mutations were still more frequent in RCC (20.1%) than in LCC (13.7%, adjusted *p* = 4.11 × 10⁻²). Overall, these findings indicate that RCC consistently exhibits higher TMB and greater enrichment of *BRAF*, *KMT2D*, and *PIK3CA* mutations, while LCC is predominantly characterized by *TP53* mutations after filtering.

**Table 2.** Comparison of TMB and Key Genetic Alterations Between LCC and RCC in non-filtered and PolyPhen/SIFT scoring-based analyses.

| Genetic alteration<br>(n, %) | LCC (n = 363) | RCC (n = 387) | Total (750) | p-value | Adjusted<br>p-value |
| --- | --- | --- | --- | --- | --- |
| **PolyPhen/SIFT score**<br>**non-filtered** |  |  |  |  |  |
| TMB (n, Mean ± SD) | 1,074 (3.45 ± 4.39) | 2,625 (6.87 ± 12.1) | 3,699 (5.88 ± 10.46) | 1.79 x 10<sup>-31</sup> | 1.61 x 10<sup>-30</sup> |
| APC | 296 (81.5) | 260 (67.2) | 556 (74.1) | 6.96 x 10<sup>-6</sup> | 1.57 x 10<sup>-5</sup> |
| TP53 | 267 (73.6) | 242 (62.5) | 509 (67.9) | 1.2 x 10<sup>-3</sup> | 1.54 x 10<sup>-3</sup> |
| KRAS | 146 (40.2) | 194 (50.1) | 340 (45.3) | 6.5 x 10<sup>-3</sup> | 7.31 x 10<sup>-3</sup> |
| PIK3CA | 76 (20.9) | 126 (32.6) | 202 (26.9) | 3.5 x 10<sup>-5</sup> | 6.3 x 10<sup>-5</sup> |
| TCF7L2 | 43 (11.8) | 51 (13.2) | 94 (12.5) | 0.582 | 0.582 |
| BRAF | 24 (6.6) | 92 (23.8) | 116 (15.5) | 8.28 x 10<sup>-11</sup> | 3.73 x 10<sup>-10</sup> |
| KMT2D | 30 (8.3) | 87 (22.5) | 117 (15.6) | 8.23 x 10<sup>-8</sup> | 2.47 x 10<sup>-7</sup> |
| SMAD4 | 38 (10.5) | 77 (19.9) | 115 (15.3) | 3.42 x 10<sup>-4</sup> | 5.13 x 10<sup>-4</sup> |
| **PolyPhen/SIFT score**<br>**filtered** | **LCC (n = 315)** | **RCC (n = 359)** | **Total (674)** | **p-value** | **Adjusted**<br>**p-value** |
| TMB (n, Mean ± SD) | 976 (3.17 ± 4.35) | 2,375 (6.65 ± 11.3) | 3,351 (5.64 ± 9.80) | 3.47 x 10<sup>-33</sup> | 3.12 x 10<sup>-32</sup> |
| TP53 | 128 (40.6) | 114 (31.8) | 242 (35.9) | 1.65 x 10<sup>-2</sup> | 2.97 x 10<sup>-2</sup> |
| KRAS | 52 (16.5) | 54 (15.0) | 106 (15.7) | 0.602 | 0.602 |
| BRAF | 21 (6.7) | 83 (23.1) | 104 (15.4) | 3.64 x 10<sup>-9</sup> | 1.64 x 10<sup>-8</sup> |
| PIK3CA | 43(13.7) | 72 (20.1) | 115 (17.1) | 2.74 x 10<sup>-2</sup> | 4.11 x 10<sup>-2</sup> |
| SMAD4 | 23 (7.3) | 47 (13.1) | 70 (10.4) | 1.39 x 10<sup>-2</sup> | 2.97 x 10<sup>-2</sup> |
| FBXW7 | 21 (6.7) | 31 (8.6) | 52 (7.7) | 0.339 | 0.381 |
| ATM | 11(3.5) | 23 (6.4) | 34 (5.0) | 0.084 | 0.108 |
| KMT2D | 10 (3.2) | 31 (8.6) | 41 (6.1) | 3.09 x 10<sup>-3</sup> | 9.27 x 10<sup>-3</sup> |

### Multivariate cox regression analysis of overall survival in colon adenocarcinoma

The multivariate Cox regression analysis for the combined cohort (n = 750) identified several significant predictors of OS (Fig. 3A). Clinical variables, including age (≥65 years; HR: 1.45, 95% CI: 1.13–1.86, *p* = 0.003), stage IV (HR: 3.06, 95% CI: 2.36–3.95, *p* < 0.001), and elevated CEA levels (>5 ng/mL; HR: 1.73, 95% CI: 1.35–2.21, *p* < 0.001), were strongly associated with worse OS. Notably, RCC was significantly associated with poorer OS compared to LCC (HR: 1.30, 95% CI: 1.02–1.66, *p* = 0.033). Among genetic factors, *BRAF* mutations emerged as a significant predictor of poor OS (HR: 1.66, 95% CI: 1.16–2.36, *p* = 0.005). Other genetic mutations in *TP53*, *KRAS*, *PIK3CA*, *SMAD4*, *FBXW7*, *ATM*, and *KMT2D*, as well as the total number of gene alterations, showed no significant association with OS.

**Figure 3.**
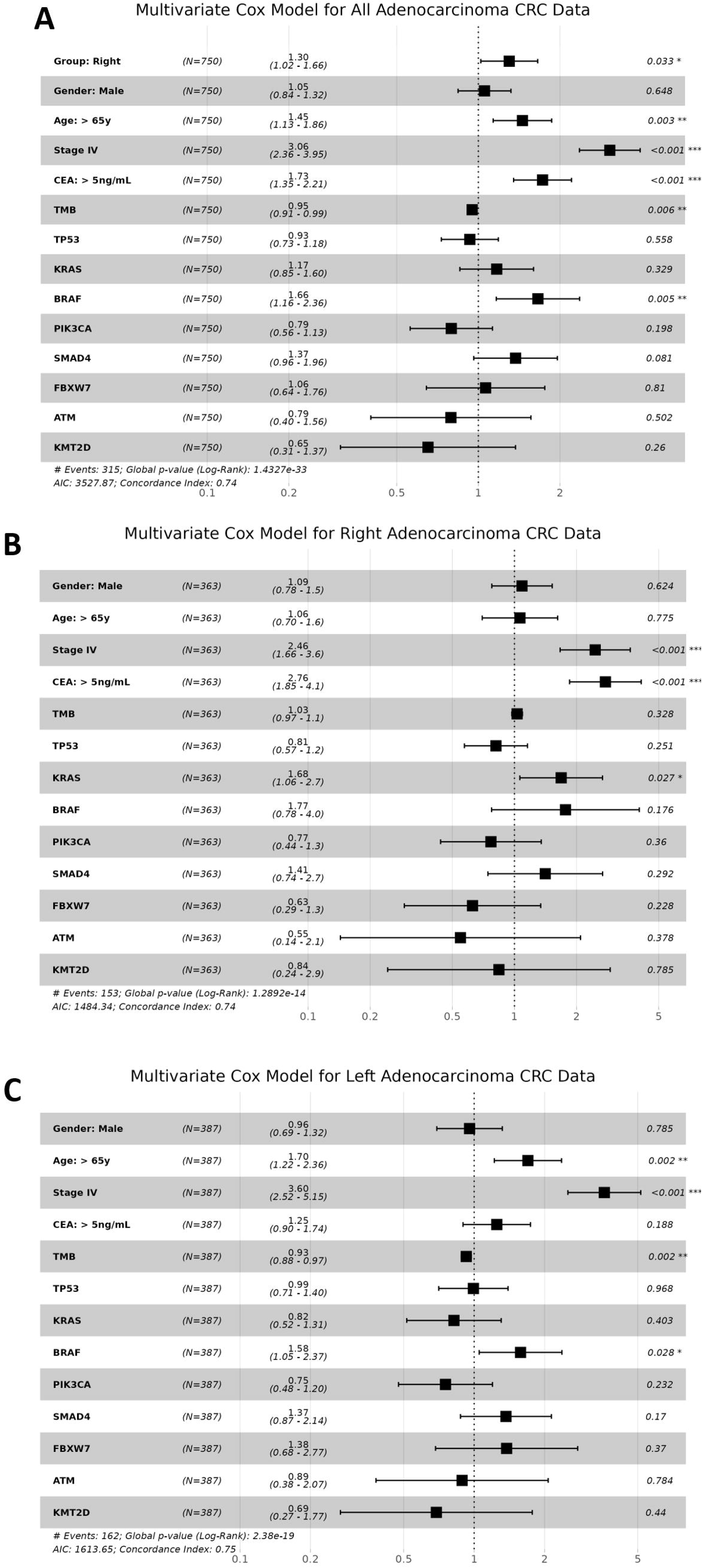
Multivariate Cox regression analysis of overall survival (OS) in colon adenocarcinoma patients. Forest plots display hazard ratios (HR) with 95% confidence intervals for clinical and genetic variables in three cohorts. (A) Combined cohort (n = 750) including both left-sided and right-sided colorectal cancers. Right-sided tumor location, age ≥65, stage IV disease, elevated CEA levels (>5 ng/mL), and *BRAF* mutations were significantly associated with worse OS. (B) Right-sided CRC cohort (RCC; n = 387), where stage IV disease, elevated CEA, and *KRAS* mutations were significantly associated with poorer outcomes. (C) Left-sided CRC cohort (LCC; n = 363), where stage IV disease, age ≥65, and BRAF mutations were associated with worse OS, while higher tumor mutation burden (TMB) was associated with improved survival. Asterisks indicate significance: *P<0.05, **P<0.01, ***P<0.001.

In the RCC group (Fig. 3B), stage IV (HR: 2.46, 95% CI: 1.66–3.6, *p* < 0.001) and elevated CEA levels (>5 ng/mL) (HR: 2.76, 95% CI: 1.85–4.1, *p* < 0.001) were the most significant predictors of poorer OS. Additionally, *KRAS* mutations were significantly associated with worse OS (HR: 1.68, 95% CI: 1.06–2.7, *p* = 0.027). Other variables, including gender, age, TMB, as well as mutations in *TP53*, *BRAF*, *PIK3CA*, *SMAD4*, *FBXW7*, *ATM*, and *KMT2D*, were not significantly associated with OS in the LCC group.

In the LCC group (Fig. 3C), stage IV (HR: 3.60, 95% CI: 2.52–5.15, *p* < 0.001) and age ≥65 years (HR: 1.70, 95% CI: 1.22–2.36, *p* < 0.001) were the strongest predictors of poorer OS. *BRAF* mutations were also significantly associated with worse OS (HR: 1.58, 95% CI: 1.05– 2.37, *p* = 0.028), while TMB showed an inverse association with OS, suggesting better outcomes with higher TMB (HR: 0.93, 95% CI: 0.88–0.97, *p* = 0.002). Other variables, including gender, CEA levels, and mutations in *TP53*, *KRAS*, *PIK3CA*, *SMAD4*, *FBXW7*, *ATM*, and *KMT2D*, showed no significant association with OS in the RCC group.

## Discussion

### Clinical and molecular differences between LCC and RCC

Using patient data from the AACR Project GENIE BPC dataset, this study confirmed the distinct characteristics of LCC and RCC, emphasizing the influence of tumor location on their molecular and clinical profiles. LCC and RCC represent two unique disease entities within the same organ, with location significantly shaping their presentation and progression. A previous comprehensive analysis of over 3,000 colon carcinoma patients, incorporating data from the PETACC3 trial and TCGA collection, demonstrated differences in their biology; revealing that RCCs frequently exhibit MSI and *BRAF*-associated signatures, while LCCs are more often characterized by chromosomal instability (13). These variations are believed to be linked to their embryological origins: the proximal colon (the cecum, ascending colon, and proximal transverse colon; RCC) develops from the midgut, whereas the distal colon (from the splenic flexure to the rectum; LCC) derives from the hindgut (14).

Through our present analysis of 750 cases of histologically confirmed adenocarcinoma, we identified significant differences in survival outcomes, mutational patterns, and genomic complexity between LCC and RCC. Our cohort revealed that, despite LCC having a significantly higher proportion of stage IV cases compared to RCC (Table. 1), patients with RCC demonstrated worse OS outcomes (Fig. 3A).

### Prognostic and survival implications of key mutations

These findings suggest that clinical features alone cannot fully explain the poorer survival outcomes observed in RCC, necessitating a deeper investigation into underlying molecular factors. To address this, we further examined genomic and clinical variables to identify independent contributors to OS. Notably, RCC exhibited significantly higher TMB (6.87 ± 12.1 vs. 3.45 ± 4.39 in LCC, adjusted *p* = 1.61 × 10⁻³⁰; Table. 2), reflecting enriched genomic instability. Previous studies have similarly reported elevated TMB in RCC compared to LCC (15, 16). This elevation is largely attributed to the prevalence of MSI-high tumors and activation of the serrated pathway (17). The serrated pathway involves precursors such as sessile serrated adenomas and serrated adenocarcinomas, which are distinct from conventional adenomas and exhibit features such as *BRAF* mutations and CIMP, commonly linked to MSI-high status (18). These molecular alterations drive genomic instability and mutation accumulation, contributing to RCC’s distinct mutational landscape.

Building on the distinct status of gene mutations in LCC and RCC, our findings further revealed significant differences in mutation patterns between the two groups. In LCC, *APC* (81.5%, adjusted *p* = 1.57 × 10⁻⁵) and *TP53* (73.6%, adjusted *p* = 1.54 × 10⁻³) mutations were highly prevalent. In contrast, RCC exhibited higher frequencies of *KRAS* (50.1%, adjusted *p* = 7.31 × 10⁻³), *PIK3CA* (32.6%, adjusted *p* = 6.3 × 10⁻⁵), *BRAF* (23.8%, adjusted *p* = 3.73 × 10⁻¹⁰), *KMT2D* (22.5%, adjusted *p* = 2.47 × 10⁻⁷), and *SMAD4* (19.9%, adjusted *p* = 5.13 × 10⁻⁴) mutations. To reduce bias and focus on identifying the most meaningful alterations, we applied standardized scoring criteria using predictive algorithms such as PolyPhen (19) and SIFT (20) scores. This approach enabled the selection of mutations with greater functional impact, allowing us to prioritize key genes in each group. After applying these criteria, *APC* mutations—initially prominent in both groups—were deprioritized as less consequential (Fig. 2B). Consequently, *TP53*, *KRAS*, and *PIK3CA* in LCC, and *TP53*, *KRAS*, *BRAF*, *PIK3CA*, and *SMAD4* were identified as the most significant mutations in RCC. Given the prominence of *TP53* and *KRAS* mutations across both groups, it is essential to first examine their shared roles before addressing the distinct pathways they influence in LCC and RCC.

In our analysis, *TP53* mutations were observed in 40.6% of LCC cases and 31.8% of RCC cases (adjusted *p* = 2.97 × 10⁻²; Table. 2), making it the most frequently mutated gene in both groups. These mutation rates, while slightly lower than those reported in some previous studies (56.0% to 73.1% in other cohorts), align with the established role of *TP53* as a key molecular driver in CC (21, 22). This slight discrepancy likely reflects the more rigorous scoring-based criteria in our study. The higher frequency of *TP53* mutations in LCC compared to RCC in our dataset mirrors trends observed in prior research, suggesting potential location-specific differences in CC (15). Additionally, survival outcomes related to *TP53* mutations differ by sidedness, with poor survival in LCC, indicating location-specific differences in the clinical implications of *TP53* mutations (23).

In this study, *KRAS* mutations were observed in 16.5% of LCC cases and 15.0% of RCC cases, with no significant difference in mutation frequency (adjusted *p* = 0.602; Table 2). These findings contrast with previous reports, where *KRAS* mutations were more frequently detected in RCC (50.1%) than in LCC (40.2%) (24, 25). Despite the little difference between groups in our cohort, *KRAS* mutations were associated with worse OS in LCC (HR: 1.64, *p* = 0.038; Fig. 3B), while no significant impact on OS was observed in RCC. This aligns with a prior study showing that *KRAS* mutations significantly worsened OS in LCC patients (HR: 1.81, 95% CI: 1.11–2.96, *p* = 0.02), while having no prognostic effect in RCC (HR: 1.03, 95% CI: 0.51–2.08, *p* = 0.95) (26). Additionally, *KRAS* mutations were found to be associated with worse 5-year OS and recurrence-free survival in LCC compared to *KRAS* wild-type (HR for OS: 1.81, 95% CI: 1.01–2.44, *p* = 0.04), while no significant survival differences were observed in RCC (HR for OS: 1.51, 95% CI: 0.73–3.14, *p* = 0.23). The consistent results across studies underscore the tumor location-dependent prognostic impact of *KRAS* mutations, suggesting that distinct biological mechanisms or co-alterations in LCC may amplify the negative survival implications of *KRAS* mutations.

*PIK3CA* mutations were observed in 13.7% of LCC cases and 20.1% of RCC cases, with a higher prevalence in RCC (adjusted *p* = 4.11 × 10⁻²; Table. 2). This corroborates previous reports that consistently demonstrated higher prevalence of *PIK3CA* mutations in RCC compared to LCC. One study reported that *PIK3CA* mutations were significantly more frequent in RCC than in LCC, particularly in tumors with poor differentiation or in advanced stages, highlighting molecular differences between groups (27). Additionally, a larger-scale analysis noted that LCC generally exhibits fewer *PIK3CA* mutations compared to RCC, further supporting the observed disparity in mutation prevalence (15).

*BRAF* mutations were significantly enriched in RCC, occurring in 23.8% of cases compared to 6.6% in LCC (Table. 2; adjusted *p* = 3.73 × 10⁻¹⁰); this finding is consistent with a prior study reporting *BRAF* mutations in 15.5% of RCC cases compared to 4.8% in LCC, further emphasizing the strong association between *BRAF* mutations and RCC (28). Additionally, in the same study, *BRAF* mutations were shown to have a significant impact on prognosis, with median OS for *BRAF*-mutant RCC being significantly worse than that for *BRAF* wild-type RCC (1,006 days vs. 1,823 days; *p* = 0.004). These results underscore the role of *BRAF* mutations in driving poor outcomes specifically in RCC. As a critical oncogene, *BRAF* mutations activate the MAPK/ERK pathway, promoting tumor progression and resistance to apoptosis, which may contribute to the worse prognosis typically observed in RCC (29). In our analysis, *BRAF* mutations emerged as the strongest predictor of unfavorable OS in the combined cohort (HR: 1.66; *p* = 0.005), with the most pronounced impact in RCC (HR: 1.61; *p* = 0.023). Similarly, another study reported that median OS for *BRAF*-mutant RCC was significantly worse than for *BRAF* wild-type RCC (480 days vs. 810 days; *p* = 0.020), consistent with prior studies that highlight its enrichment in RCC (30).

In this study, *SMAD4* mutations were detected in 7.3% of LCC cases and 13.1% of RCC cases, demonstrating a significantly higher prevalence in RCC (adjusted *p* = 2.97 × 10⁻²; Table. 2). This enrichment of *SMAD4* mutations in RCC aligns with its known role in promoting proximal tumor progression and metastasis. Similarly, a prior large-scale study of 1,876 patients with CC reported *SMAD4* mutation rates of 16% in RCC compared to 10% in LCC (*p* = 0.0020) (15). Another study also observed *SMAD4* mutations in 19.8% of CC cases, with higher prevalence in RCC than in LCC (31). Prior research has shown that *SMAD4* mutations in RCC are associated not only with their higher prevalence but also with worse survival outcomes and higher TMB compared to LCC, further emphasizing their impact on the aggressive nature of RCC (32).

This study had several limitations. First, as a secondary analysis of the GENIE BPC dataset, the analysis was inherently constrained by incomplete or missing clinical and/or genomic data. Key variables, such as specific treatment regimens, performance status, or MSI/MMR status, were not consistently available, limiting the depth of analysis for certain associations. While we compared treatment regimens between LCC and RCC across both stage I–III (Table. S1) and stage IV (Table. S2) cohorts, no statistically significant differences were observed. This may reflect a lack of sufficient statistical power to detect differences or the possibility that treatment strategies were relatively uniform within these groups, limiting our ability to evaluate the differential impact of therapies based on tumor location. Second, genomic sequencing panels used across contributing institutions differed in gene coverage, potentially impacting the detection of less prevalent mutations. Although we accounted for this by restricting analyses to genes included in the sequencing panels, variability in sequencing methodologies may still introduce biases in the identification of mutations. Third, locoregional treatments, such as radiation or surgery, and their potential influence on survival outcomes, were not captured, further limiting the ability to fully contextualize the observed associations. Lastly, we retrospectively analyzed the associations between tumor location, genetic alterations, and prognosis. Prospective validation and more comprehensive datasets are required for robust assessments.

In conclusion, the present findings highlight the clinical and molecular landscapes of LCC and RCC, underscoring the importance of both tumor location and key genetic alterations in shaping patient outcomes. *BRAF* emerged as a critical driver of poor prognosis in RCC, while *KRAS* mutations were closely tied to worse outcomes in LCC. Despite these evident associations, the persistent survival gaps between LCC and RCC indicate that additional molecular and clinical factors likely influence disease progression. Recognizing these complexities warrants prospective studies with diverse patient populations to refine our understanding of CC heterogeneity and further clarify tumor progression mechanisms.

## Data Availability

All data produced in the present work are contained in the manuscript

## Acknowledgements

This research utilized data generated by the AACR Project GENIE consortium. The authors recognize the invaluable efforts of all consortium members who have advanced collaborative data sharing in cancer research. We further acknowledge the support provided by the AACR, including its financial and infrastructural contributions.

## Funding

This work was supported by Dankook University.

## Availability of data and materials

In alignment with the principles of open science, the dataset supporting this study’s conclusions is available in the AACR Project GENIE repository, hosted on the Synapse platform. The GENIE BPC CRC 2.0-public data release is available for download at https://www.synapse.org/Synapse:syn27056172/wiki/616601. Data access requires registration and compliance with the dataset-specific terms of use, ensuring ethical usage and broad dissemination of the findings.

## Authors’ contributions

All authors listed have made a substantial, direct, and intellectual contribution to the work and approved it for publication.

## Ethics approval and consent to participate

Not applicable.

## Patient consent for publication

Not applicable.

## Competing interests

The authors declare that the research was conducted in the absence of any commercial or financial relationships that could be construed as a potential conflict of interest.

## Notes

### Competing Interest Statement

The authors have declared no competing interest.

### Author Declarations

https://www.synapse.org/Synapse:syn27056172/wiki/616601.

