## Supplemental Tables for "A Comparative Genomic Analysis of Left- and Right-Sided Colon Cancer Using Real-World Data from the AACR Project GENIE BPC Dataset"

**Supplementary Table S1.** Treatment regimens for stage I–III colon cancer patients, stratified by LCC and RCC, including targeted therapies and chemotherapy across different treatment lines. The table reports patient distributions and p-values for group comparisons.

| Regimen Type | 1st line | 2nd line | 3rd line | *p*-value |
|---|---|---|---|---|
| **Bevacizumab +** | | | | 0.301 |
| Left | 8 | 29 | 31 | |
| Right | 11 | 38 | 24 | |
| **Cetuximab +** | | | | 0.865 |
| Left | 0 | 2 | 6 | |
| Right | 0 | 3 | 5 | |
| **Panitumumab +** | | | | 0.389 |
| Left | 1 | 2 | 2 | |
| Right | 0 | 1 | 5 | |
| **ICI** | | | | |
| Left | 2 | 0 | 0 | |
| Right | 1 | 2 | 4 | |
| **Chemotherapy** | | | | |
| Left | 131 | 65 | 35 | |
| Right | 159 | 72 | 52 | |
| **N/A** | | | | |
| Left | 19 | 63 | 87 | |
| Right | 46 | 101 | 127 | |

**Supplementary Table S2.** Treatment regimens for stage IV colon cancer patients, stratified by LCC and RCC, including targeted therapies and chemotherapy across different treatment lines. The table presents patient distributions and p-values for group comparisons.

| Regimen Type | 1st line | 2nd line | 3rd line | *p*-value |
|---|---|---|---|---|
| **Bevacizumab +** | | | | 0.278817 |
| Left | 69 | 79 | 59 | |
| Right | 66 | 56 | 38 | |
| **Cetuximab +** | | | | 0.677764 |
| Left | 2 | 5 | 9 | |
| Right | 0 | 1 | 5 | |
| **Panitumumab +** | | | | 0.501081 |
| Left | 3 | 4 | 13 | |
| Right | 3 | 1 | 5 | |
| **ICI** | | | | |
| Left | 0 | 0 | 0 | |
| Right | 0 | 2 | 5 | |
| **Chemotherapy** | | | | |
| Left | 121 | 88 | 76 | |
| Right | 94 | 83 | 59 | |
| **N/A** | | | | |
| Left | 7 | 26 | 45 | |
| Right | 7 | 27 | 58 | |
